# Lack of ownership of mobile phones could hinder the rollout of mHealth interventions in Africa

**DOI:** 10.1101/2022.05.10.22274883

**Authors:** Justin T. Okano, Joan Ponce, Matthias Krönke, Sally Blower

## Abstract

**Background:** Mobile Health interventions, which require ownership of mobile phones, are being investigated throughout Africa. We estimate the percentage of individuals who own mobile phones in 33 African countries, identify a relationship between ownership and proximity to a health clinic (HC), and quantify inequities in ownership. We investigate basic mobile phone (BPs) and smartphones (SPs): SPs can connect to the internet, BPs cannot.

**Methods:** We use nationally representative data collected in 2017—2018 from 45,823 individuals in Round 7 of the Afrobarometer surveys. We use Bayesian Multilevel Logistic regression models for our analyses.

**Results:** 82% of individuals in the 33 countries own mobile phones: 42% BPs, 40% SPs. Individuals who live close to an HC have higher odds of ownership than those who do not (adjusted odds ratio [aOR]: 1.31, Bayesian 95% Highest Posterior Density [HPD] region: 1.24— 1.39). Men, compared with women, have over twice the odds of ownership (aOR: 2.37, 95% HPD region: 1.96—2.84). Urban residents, compared with rural residents, have almost three times the odds (aOR: 2.66, 95% HPD region: 2.22—3.18) and, amongst mobile phone owners, nearly three times the odds of owning an SP (aOR: 2.67, 95% HPD region: 2.33—3.10). Ownership increases with age, peaks in 26—40 year olds, then decreases. Individuals under 30 are more likely to own an SP than a BP, older individuals more likely to own a BP than an SP. Probability of ownership decreases with the Lived Poverty Index; however, some of the poorest individuals own SPs.

**Conclusions:** If the digital devices needed for mHealth interventions are not equally available within the population (which we have found is the current situation), rolling out mHealth interventions in Africa is likely to propagate already existing inequities in access to healthcare.

**Funding:** National Institute of Allergy and Infectious Diseases

## INTRODUCTION

The effectiveness of mobile Health (mHealth) based interventions are currently under investigation in many countries in Africa (Ag Ahmed et al., 2017; Manby et al., 2022; Onukwugha et al., 2022; Osei et al., 2020). The objective of these interventions is to increase access to healthcare resources. Such interventions, if shown to be cost-effective, would be extremely important in Africa where many people need to travel long distances to reach healthcare facilities (World Health Organization, 2021a). However, in order to use mHealth interventions, individuals need to own a mobile phone; furthermore, many proposed interventions require ownership of smartphones. In the continent of Africa mobile phone ownership has been reported to have increased rapidly over the past decade (Krönke, 2020), but country-level differences in ownership have not been quantified, nor have country-level inequities in ownership been compared. Here we address these knowledge gaps for 33, out of the 54, countries in Africa: together, these 33 countries encompass ∼60% of the population of the African continent, which contains ∼1.4 billion people. Specifically, we: (i) estimate the percentage of individuals who own mobile phones in all 33 countries together, and for each country at the national- and sub-national level, (ii) identify a relationship between ownership of a mobile phone and proximity to a health clinic (HC), and (iii) identify inequalities/inequities in the ownership of mobile phones based on gender, urban-rural residency, age, and poverty. We investigate the ownership of a basic mobile phone (BP; a mobile phone that cannot connect to the internet) and a smartphone (SP; a mobile phone that can connect to the internet) separately. Finally, we discuss the implications of our results for designing and implementing mHealth interventions in Africa.

To conduct our analyses, we used nationally representative sample data collected from 45,823 individuals in 33 countries in Round 7 (R7) of the Afrobarometer (Afrobarometer, 2021) survey: R7 was conducted in 2017—2018. Afrobarometer is a pan-African, non-partisan research project that has been operating since 1999. It is the world’s leading source of high-quality public opinion data for Africa. The surveys measure citizen’s attitudes on democracy, governance, society as well as the economy, and the continent’s progress towards achieving the UN Sustainable Development Goals (Brass et al., 2019; Krönke et al., 2022; Mattes, 2019). The surveys also collect data on the ownership of mobile phones. Further details on the Afrobarometer data are given in the Methods.

Problems due to the geographic inaccessibility of healthcare in Africa have been well documented (World Health Organization, 2021a). These problems reflect the resource constraints on the healthcare system, and hence are more acute in some countries in Africa than others: e.g., Botswana is a middle-income country with a well-financed healthcare system, and Malawi is one of the poorest countries in the world with a healthcare system that is severely financially constrained. However, in all African countries, the problem of geographic inaccessibility to healthcare is particularly pronounced in rural areas, where many of the poorest citizens live. In rural areas, the problem of the need to travel long distances to reach HCs is exacerbated by the lack of transportation: many individuals in rural areas have to walk to reach HCs (Palk et al., 2020; World Health Organization, 2021a). The phenomenon of distance decay in utilization of HCs has been observed in many African countries: the further individuals live from an HC, the less likely they are to utilize healthcare (Lankowski et al., 2014). The geographic inaccessibility of HCs has been shown to be associated with decreased utilization of antenatal care and bed nets (for protection against malaria), lower vaccination rates, higher attrition rates from HIV and TB treatment programs, lower adherence levels in HIV programs, and reduced maternal fever-seeking behavior, and uptake in contraception (Alegana et al., 2018; Bilinski et al., 2017; Ouma et al., 2017; Terzian et al., 2018).

Currently there are multiple mHealth interventions that are being investigated or being used at a small-scale in almost every African country; the number of mHealth interventions is continuing to increase. Throughout the continent, mHealth interventions are being used for multiple reasons: for disease diagnosis and treatment support by health workers (Anstey Watkins et al., 2018), to increase adolescent’s use of sexual and reproductive health services (Onukwugha et al., 2022), for HIV prevention and management (Manby et al., 2022), to improve maternal and child health (Ag Ahmed et al., 2017), to support the COVID-19 response (Fischer et al., 2021), and to improve surveillance coverage for new outbreaks of infectious diseases (e.g., Ebola) (Tom-Aba et al., 2018).

## METHODS

The Afrobarometer R7 survey (Afrobarometer, 2021) collected data in 34 African countries: Benin, Botswana, Burkina Faso, Cabo Verde, Cameroon, Cote d’Ivoire, Eswatini, Gabon, Gambia, Ghana, Guinea, Kenya, Lesotho, Liberia, Madagascar, Malawi, Mali, Mauritius, Morocco, Mozambique, Namibia, Niger, Nigeria, Sao Tome and Principe, Senegal, Sierra Leone, South Africa, Sudan, Tanzania, Togo, Tunisia, Uganda, Zambia, and Zimbabwe. We excluded Kenya from our analysis as its questionnaire did not differentiate between SP and BP ownership.

We used the Afrobarometer data (Afrobarometer, 2021) to estimate the probability of owning a mobile phone (either a BP or an SP) and, amongst mobile phone owners, the probability of owning an SP. We made these estimates at three levels: (i) multi-country (aggregating data from all 33 countries), (ii) the national-level for each country, and (iii) the sub-national level within each country. To make these estimates, we used data from the *N* = 43,925 individuals in the 33 countries who provided data on mobile phone ownership: i.e., on whether they owned one (*n* = 35,654), didn’t own one (*n* = 4,895), or didn’t own one but someone in their house owned one (*n* = 3,376). Mobile phone ownership was recoded as a binary variable; participants who reported that someone else in their household owned a mobile phone were coded as not owning a phone. Contingent on mobile phone ownership, participants were asked whether or not their phone had internet access; individuals who answered in the affirmative were coded as owning an SP (*n* = 16,812); individuals who answered ‘don’t know’ or refused to answer were excluded (*n* = 252); the remaining participants were coded as owning a BP (*n* = 18,590). Afrobarometer ‘within-country’ weights were used for all national-level estimates (The Afrobarometer Network, 2021), multi-country estimates were made by weighting the national-level estimates with UN population data (United Nations, 2019). Sub-national estimates of mobile phone ownership were mapped by linking current national and sub-national boundaries (GADM, 2021) to Afrobarometer data at the province/state level.

To identify inequalities/inequities in ownership we used data on five variables from the Afrobarometer R7 survey: gender, age, poverty, urban/rural residency, and proximity to an HC (Afrobarometer, 2021). We defined poverty, as in the Afrobarometer surveys, by using the Lived Poverty Index (LPI). This index is calculated by combining answers to five survey questions that measure how often individuals have gone without basic necessities such as water, food, and medical care in the past month (Mattes, 2020). We use a four-point scale for LPI, where 0 indicates an individual is in the wealthiest group in terms of accessing basic necessities, and 3 indicates an individual is in the poorest group. We defined proximity to an HC as a binary variable: close, or not. Individuals who had an HC present in the enumeration area of their residence, or within easy walking distance thereof, were considered to be in close proximity to an HC. We estimated country-specific Crude Odds Ratios (cORs) of mobile phone ownership separately by gender and urban-rural status, calculated an age- and gender-stratified population-pyramid of mobile phone ownership, and constructed Bayesian models for (i) the ownership of mobile phones and (ii) the ownership of SPs amongst mobile phone owners.

To specify the Bayesian logistic regression (BLR) models for the probability of owning a mobile phone, we modeled phone ownership *y*_*ij*_ of individual *i* ∈ (1, *n*_*j*_) in country *j* ∈ (1,33) as a Bernoulli variable with probability ***θ***_*ij*_:

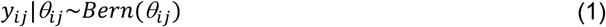

The probability of owning a mobile phone, *P*(*y*_*ij*_ = 1) = ***θ***_*ij*_, was then modeled using the logit-link function:

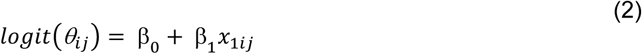

where β_0_ is the population-level intercept, and β_1_ is a regression coefficient that quantifies the influence of predictor variable *x*_1*ij*_; we used a separate BLR model for each of the five variables. Notably, 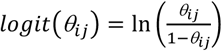 is the log-odds of mobile phone ownership. The BLR models for the probability, for mobile phone owners, of owning an SP are defined equivalently, with *y*_*ij*_ now representing ownership of a SP by phone owner *i* in country *j*.

We then constructed Bayesian multilevel logistic regression (BMLR) models (Gelman et al., 2013) for the probability of owning a mobile phone (model 1), and the probability, for mobile phone owners, of owning an SP (model 2). These models enabled us to quantify the effect of each of the five variables whilst accounting for the effect of the other four variables and the nested structure of the data. We specified model 1 by modifying equation (2):

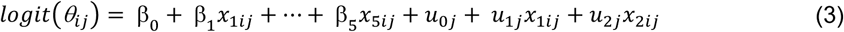

Here the five predictor variables are denoted by *x*_*kij*_ and their associated regression coefficients by *β*_*k*_ (*k* ∈ (1,5)). 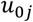 are country-level intercepts, 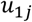 are coefficients for the country-level effect of urban/rural residency *x*_1*ij*_, and 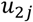 are coefficients for the country-level effect of gender *x*_2*ij*_. The country-level effects are distributed as Multivariate Normal with mean 0 and unstructured covariance matrix ∑:

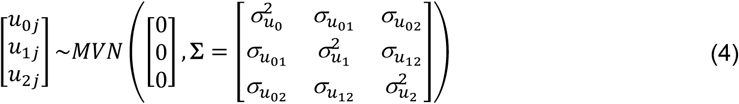

By substituting 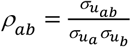 (where ρ_*ab*_ is the correlation between country-level effects *u*_*a*_ and *u*_*b*_), **∑** can be reparametrized as a function of the correlation matrix (***R***). We derive this for model 1; the result is generalizable.

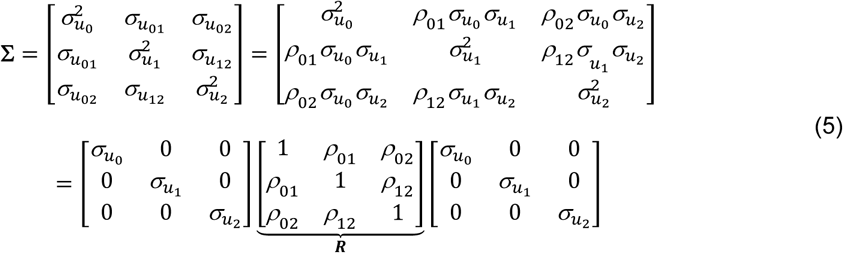

Model 2 for SP ownership (amongst mobile phone owners) is defined in an equivalent manner, with the addition of interaction effects between gender and proximity to an HC. This allows us to discern differences in SP ownership between women who do not live in close proximity to an HC and: i) women who live in close proximity to an HC, ii) men who do not live in close proximity to an HC, and iii) men who live in close proximity to an HC.

We used weakly informative priors (Gelman, 2006; Gelman et al., 2013; Gelman et al., 2008; Lemoine, 2019; Williams et al., 2018):

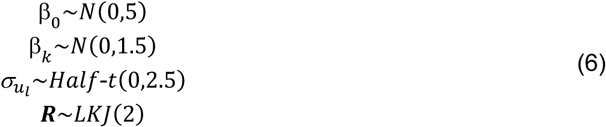

For the population-level effects **β**_*k*_ *k* ∈ (1, *N*), we chose Normal priors with mean 0 and standard deviation 1.5. This necessitates the initial sampling to not exclude effects as large as ±3.9; i.e., the 99% bound for this distribution. This corresponds to being able to detect an odds ratio as large as 48, or as small as 1/48. We used similar reasoning to define the other priors. We used half-t priors for the standard deviation parameters (Gelman, 2006) to ensure positivity. Finally, we used an LKJ(2) prior for the correlation matrix ***R***; this distribution (Lewandowski et al., 2009) is bounded by [-1,1] and centered at 0 with edge values less likely.

We fit the BLR and BMLR models by using Markov Chain Monte Carlo (MCMC) sampling to approximate the posterior distributions of all model parameters. MCMC sampling was conducted in the programming language Stan (Stan Development Team, 2020) via the R package brms (v. 2.15.0) (Bürkner, 2017). 10,000 posterior samples were drawn over 4 chains, following a warm-up of 1,000 for each. Standard MCMC diagnostics (Betancourt, 2017; Gelman et al., 2013; Gelman & Rubin, 1992) were used to assess chain convergence, independence, and sampling efficiency. We made density plots of each parameter’s posterior distribution: medians and Bayesian 95% Highest Posterior Density (HPD) regions. For all five variables, we computed median ORs (cORs for the BLR models, and Adjusted Odds Ratios, aORs, for the BMLR models) and the corresponding 95% HPD regions. Receiver Operating Characteristic (ROC) curves were used to assess the diagnostic capabilities of the fitted BMLR models.

## RESULTS

We found that 82% of individuals in the 33 countries own mobile phones; 42% of individuals own BPs, 40% own SPs. Figure 1A shows the probability of phone ownership in all 33 countries. Overall, the probability of not owning a mobile phone (either a BP or a SP) is fairly low (median: 0.15), but there is substantial variation between countries: the probability ranges from 0.05 to 0.53 with a standard deviation (*s*) of 0.12. The probability of owning a BP is moderately high (median: 0.40), but again – there is substantial variation amongst countries, ranging from 0.23 to 0.66 with *s* = 0.10. Overall, the country-level probability of owning a BP is not significantly different (t = 0.76; *P* = 0.45) from the probability of owning an SP, but the probability of owning an SP is much more variable amongst countries (*s* = 0.16). Figure 1B shows the country-specific probabilities of ownership of a BP (orange data) or an SP (red data). Notably, there is substantial sub-national variation, within almost every one of the 33 countries, in the ownership of a BP (Figure 1C) or of an SP (Figure 1D). Even in countries, such as South Africa, where there is a high probability of owning a mobile phone, there is a very low probability of ownership in certain areas of the country.

**Figure 1:**
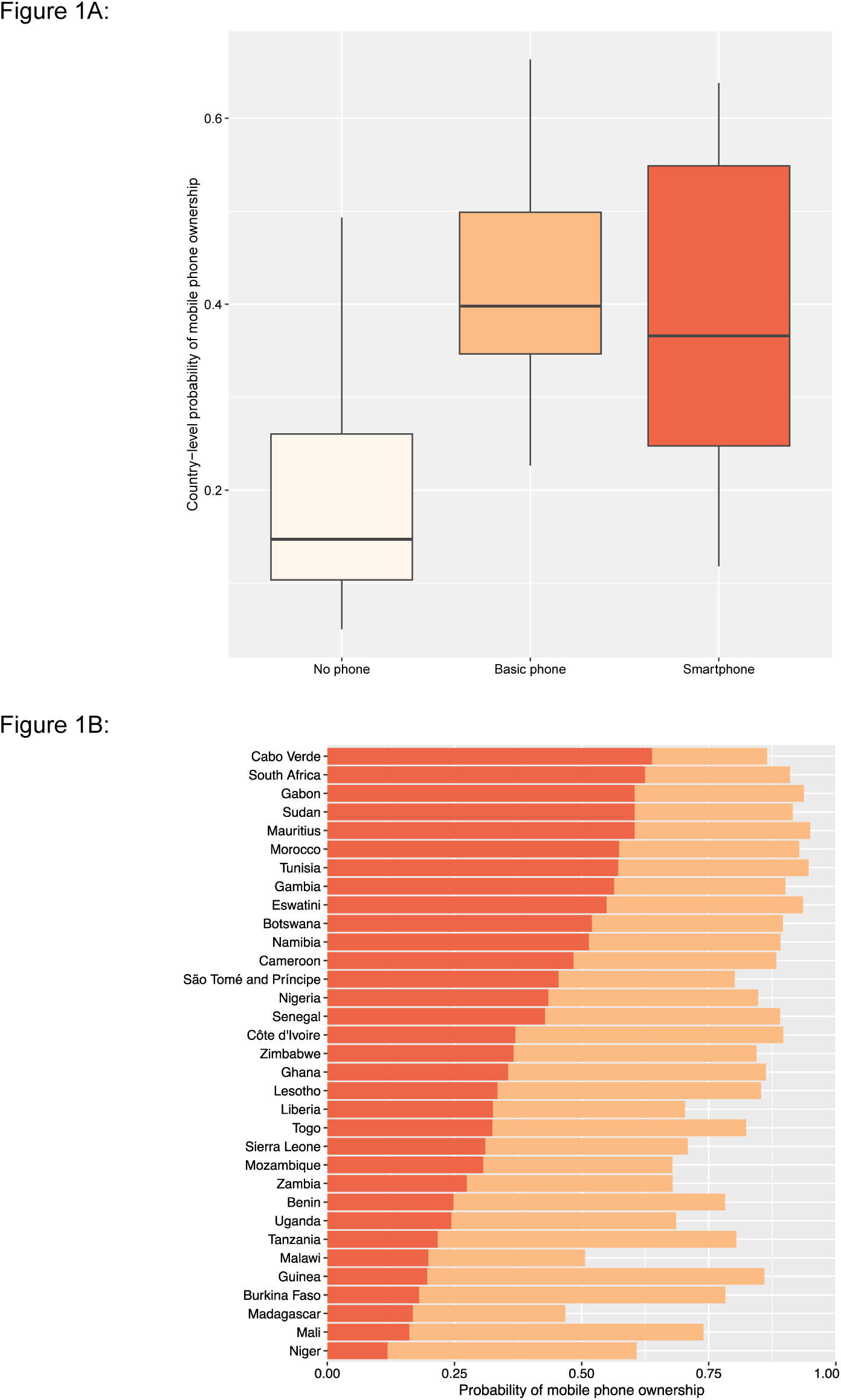
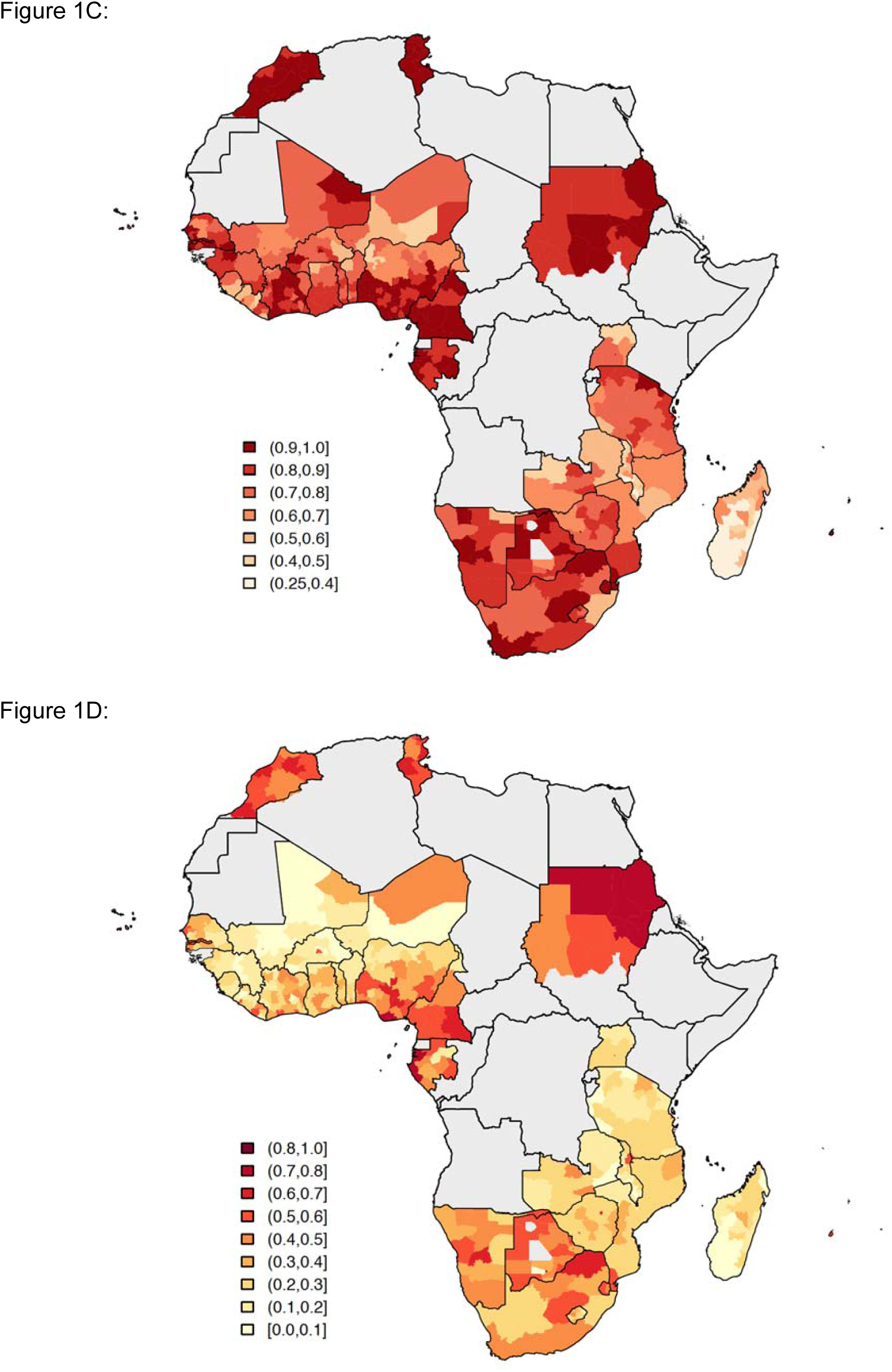
Basic mobile phone and smartphone ownership in 33 African countries. **(A)** Boxplots show the probabilities of not owning a mobile phone (beige), owning a BP (orange), or owning an SP (red). Country-level probabilities (dots) are overlaid and jittered to reduce overlap. **(B)** Barplot shows the country-level probabilities of BP ownership (orange) and SP ownership (red) ordered by SP ownership. Geographic distribution showing probabilities of (**C)** BP ownership and (**D)** SP ownership in 33 Afrobarometer countries at the sub-national level.

We found a very clear relationship between country-level ownership of a mobile phone and living in close proximity to an HC (Figure 2A). SP owners were more likely than BP owners to live close to an HC (t = 6.88; *P* < 0.001); in turn, BP owners were significantly more likely than individuals who did not own a mobile phone to live close to an HC (t = 5.50; *P* < 0.001). Figure 2B shows, for each of the 33 countries, the proportion of individuals who live in close proximity to an HC based on whether they own an SP, own a BP, or do not own a mobile phone. Again, there is considerable variation amongst countries.

**Figure 2:**
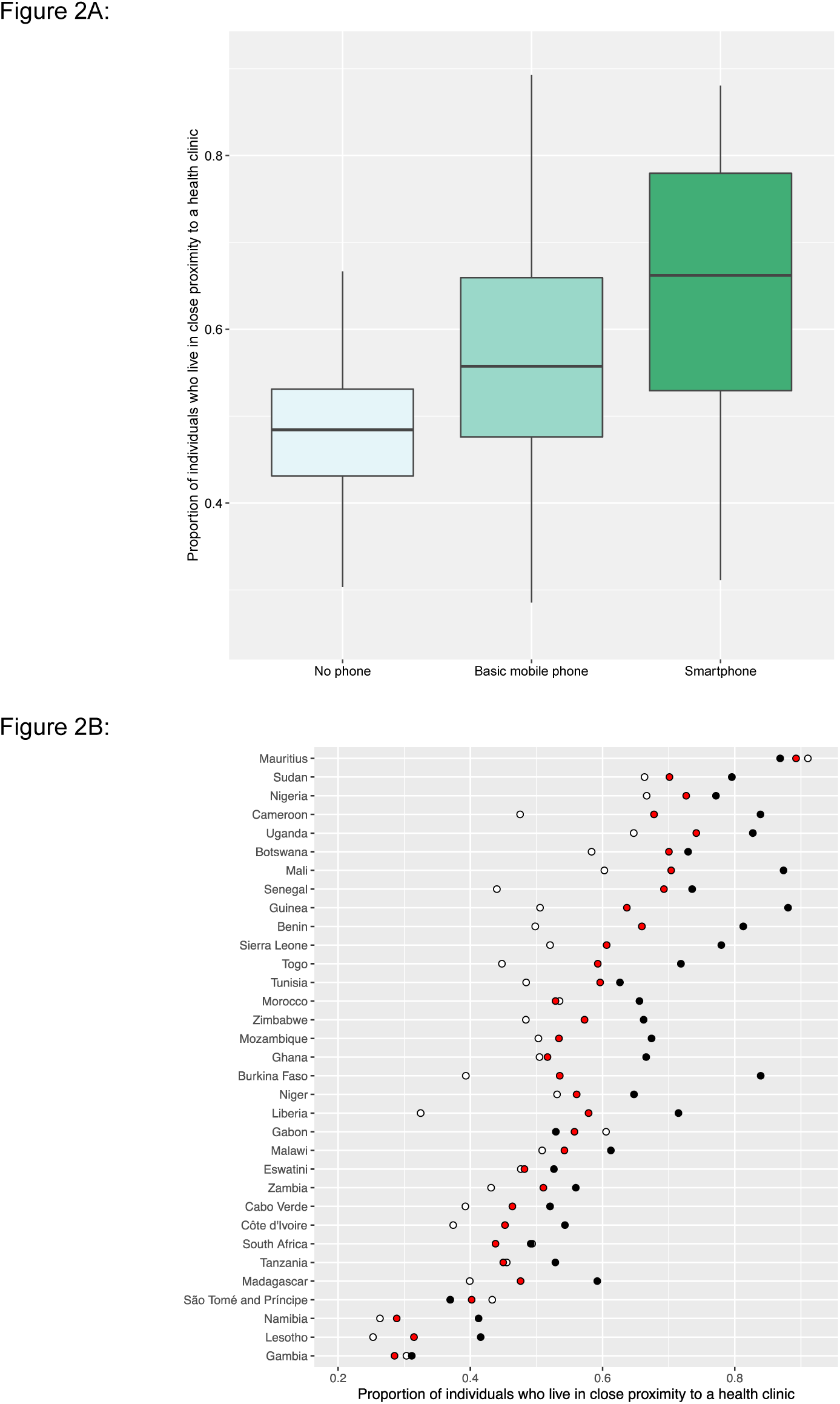
Proximity to health clinics and mobile phone ownership in 33 African countries. (**A)** Boxplots show the proportion of individuals who live in close proximity to a HC based on whether they do not own a mobile phone (mean 0.48), own a BP (mean 0.56), or own an SP (mean 0.66). Country-level probabilities (dots) are overlaid and jittered to reduce overlap. (**B)** Scatterplot shows the country-specific proportions of individuals who live in close proximity to a HC amongst individuals who: do not own a mobile phone (white dots), own a BP (red dots), or own an SP (black dots).

Considering the 33 countries together, we found a gender inequity in ownership of mobile phones: 87% of men versus 76% of women own mobile phones. We also found a gender inequity in ownership of SPs: 50% of men (who own mobile phones) own SPs versus 44% of women (who own mobile phones). We found that men are significantly more likely than women to own mobile phones in 27 out of the 33 countries (Table 1): the OR of male to female phone ownership is greatest in Mali and Benin (cOR 4.77). We did not find a significant gender difference in Botswana, Cabo Verde, Gabon, Lesotho, Namibia, or South Africa. Amongst mobile phone owners, men are more likely than women to own SPs in 15 of the 33 countries (Table 1): gender inequity is most pronounced in Benin (cOR 2.85).

**TABLE 1:** Probability of mobile phone ownership by country and gender

| Country | Any mobile phone |  |  | Smartphone |  |  |
| --- | --- | --- | --- | --- | --- | --- |
|  | Female | Male | OR<sup>†</sup> | Female | Male | OR<sup>†</sup> |
| Mali | 0.60 | 0.88 | 4.77* | 0.15 | 0.26 | 1.93* |
| Benin | 0.66 | 0.90 | 4.77* | 0.19 | 0.41 | 2.85* |
| Burkina Faso | 0.67 | 0.90 | 4.31* | 0.18 | 0.27 | 1.64* |
| Senegal | 0.83 | 0.95 | 3.90* | 0.44 | 0.52 | 1.40* |
| Guinea | 0.79 | 0.93 | 3.46* | 0.22 | 0.23 | 1.06 |
| Niger | 0.47 | 0.74 | 3.21* | 0.14 | 0.23 | 1.92* |
| Cote d'Ivoire | 0.85 | 0.95 | 3.08* | 0.33 | 0.49 | 1.97* |
| Nigeria | 0.78 | 0.91 | 3.00* | 0.48 | 0.54 | 1.29* |
| Togo | 0.75 | 0.89 | 2.80* | 0.33 | 0.45 | 1.61* |
| Gambia | 0.86 | 0.94 | 2.78* | 0.60 | 0.65 | 1.23 |
| Uganda | 0.59 | 0.80 | 2.76* | 0.33 | 0.37 | 1.18 |
| Tunisia | 0.92 | 0.97 | 2.45* | 0.61 | 0.60 | 0.97 |
| Sierra Leone | 0.62 | 0.80 | 2.42* | 0.44 | 0.44 | 1.01 |
| Tanzania | 0.74 | 0.87 | 2.41* | 0.23 | 0.31 | 1.50* |
| Malawi | 0.41 | 0.61 | 2.29* | 0.34 | 0.43 | 1.49* |
| Morocco | 0.90 | 0.95 | 2.27* | 0.59 | 0.64 | 1.21 |
| Ghana | 0.82 | 0.91 | 2.14* | 0.35 | 0.47 | 1.70* |
| Mauritius | 0.93 | 0.97 | 2.11* | 0.63 | 0.64 | 1.02 |
| Cameroon | 0.85 | 0.92 | 2.09* | 0.55 | 0.54 | 0.96 |
| Mozambique | 0.62 | 0.75 | 1.86* | 0.45 | 0.45 | 1.02 |
| Sao Tome and Principe | 0.75 | 0.85 | 1.83* | 0.55 | 0.58 | 1.14 |
| Liberia | 0.64 | 0.76 | 1.79* | 0.38 | 0.53 | 1.90* |
| Sudan | 0.89 | 0.94 | 1.77* | 0.65 | 0.67 | 1.11 |
| Eswatini | 0.92 | 0.95 | 1.74* | 0.57 | 0.60 | 1.14 |
| Zambia | 0.63 | 0.73 | 1.61* | 0.40 | 0.41 | 1.06 |
| Zimbabwe | 0.82 | 0.87 | 1.46* | 0.42 | 0.45 | 1.15 |
| Madagascar | 0.44 | 0.50 | 1.28* | 0.36 | 0.36 | 0.99 |
| Gabon | 0.93 | 0.94 | 1.26 | 0.63 | 0.66 | 1.13 |
| Namibia | 0.88 | 0.90 | 1.24 | 0.54 | 0.62 | 1.40* |
| Botswana | 0.89 | 0.91 | 1.23 | 0.55 | 0.62 | 1.33* |
| Cabo Verde | 0.85 | 0.88 | 1.20 | 0.71 | 0.77 | 1.37* |
| Lesotho | 0.84 | 0.86 | 1.17 | 0.39 | 0.40 | 1.03 |
| South Africa | 0.91 | 0.91 | 0.97 | 0.69 | 0.68 | 0.93 |

\*Significant at $\alpha=0.05$ <sup>†</sup>Odds ratio of male ownership to female ownership

Considering the 33 countries together, we found a substantial inequity in ownership of mobile phones based on whether individuals lived in urban or rural areas: 91% of urban residents versus 74% of rural residents. We also found an urban-rural inequity in ownership of SPs: 61% of urban residents (who own mobile phones) own SPs versus 34% of rural residents (who own mobile phones). We found significant urban-rural differences in the ownership of mobile phones in 29 of the 33 countries (Table 2): the OR of urban to rural phone ownership is greatest in Gabon (cOR 7.17). Notably, we did not find a significant urban-rural inequity in Eswatini, the Gambia, Mauritius, or South Africa. Urban residents are more likely, than rural residents, to own SPs in 31 of 33 countries (Table 2); the urban/rural difference in SP ownership is greatest in Burkina Faso (cOR 7.57). We did not find significant urban-rural differences in ownership of SPs in the Gambia or Sao Tome and Principe.

**TABLE 2:**
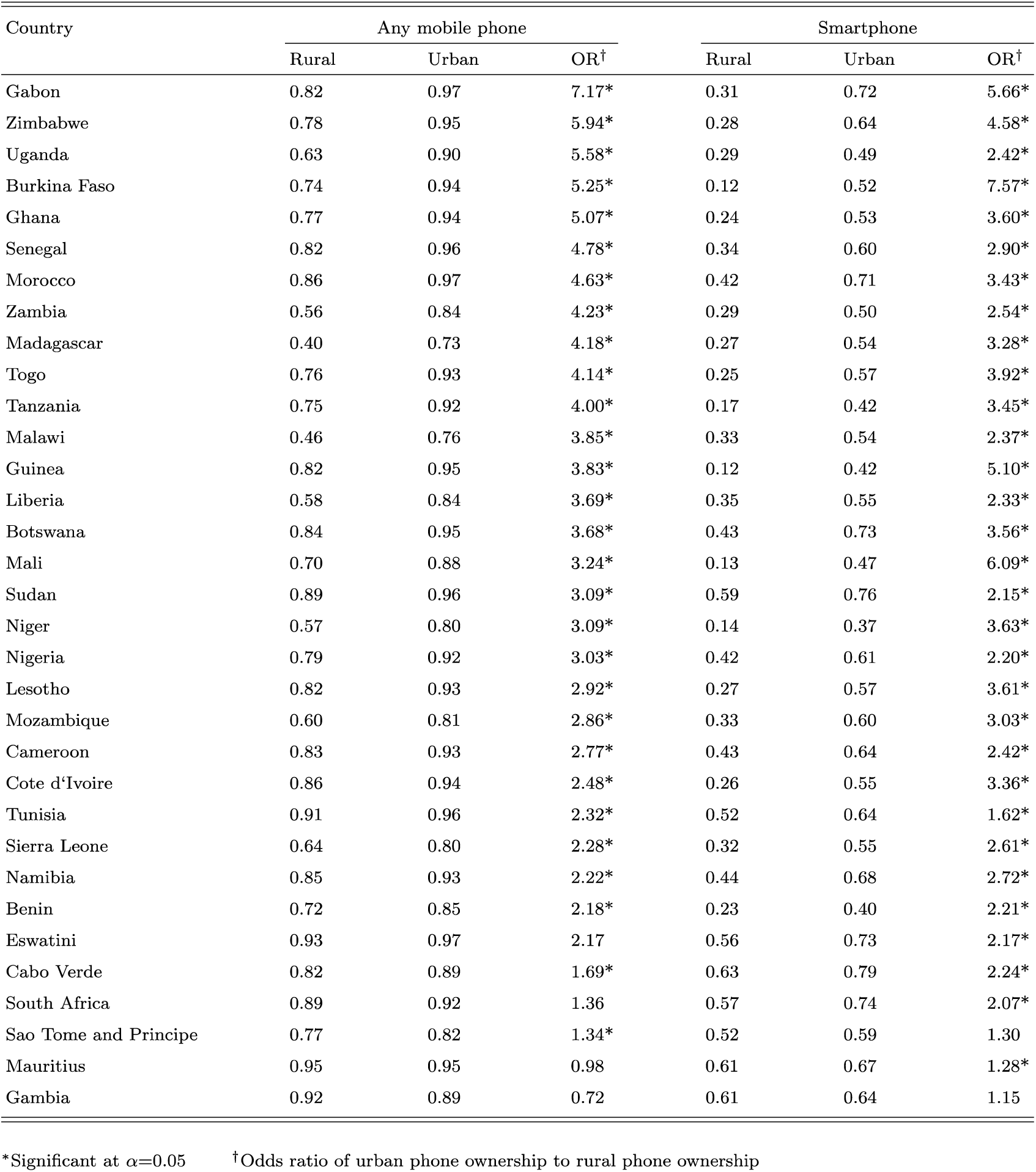
Probability of mobile phone ownership by country and urban/rural status

The age and gender-stratified ownership pyramid, shown in Figure 3, is based upon aggregated data from all 33 countries. The age pyramid shows that ownership of mobile phones differs substantially amongst age classes, by type of mobile phone (BP or SP), and by gender. In all age classes, a high proportion of individuals own mobile phones; almost half of phone owners are under 30 years old. For women, 18—30 year olds (and for men, 18—35 year olds) are more likely to own an SP than a BP; whereas, the opposite holds true in the older age classes. In all age classes, a greater proportion of men than women own mobile phones; this gender inequity in ownership is accentuated for SPs.

**Figure 3:**
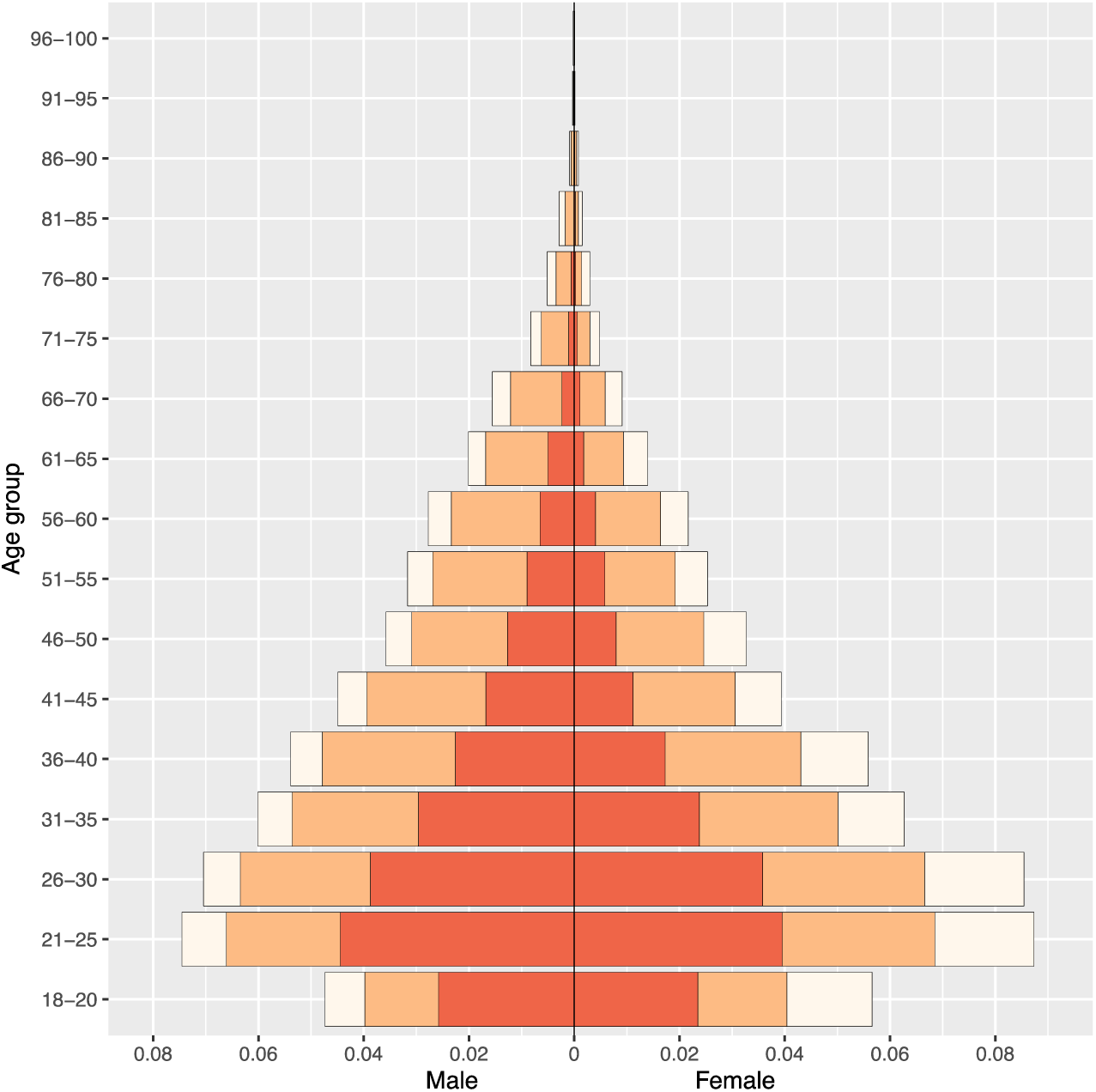
Phone ownership by age and gender in 33 African countries. Population pyramid displays the distribution of the population stratified by gender and 5-year age groupings (with the exception of the 18-20 age class) by ownership of a mobile phone: no mobile phone (tan), a BP (orange), or an SP (red).

Results from the BLR and BMLR models on the probability of owning a mobile phone are shown in Table 3. All ORs are significantly different from 1 in both the bivariable and multivariable analysis. Men have over twice the odds of owning a mobile phone than women (aOR: 2.37, 95% HPD region: 1.96—2.84). Urban residents have nearly three times the odds of owning a mobile phone than rural residents (aOR: 2.66, 95% HPD region: 2.22—3.18). Ownership of mobile phones increases with age, peaks in 26 to 40 year olds, and then decreases. The probability of ownership decreases with the LPI: the wealthiest individuals have approximately three times higher odds of owning mobile phones than the poorest individuals (aOR: 2.87, 95% HPD region: 2.53—3.27). Notably, individuals who live in close proximity to an HC have higher odds of owning mobile phones than individuals who do not live in close proximity to an HC (aOR: 1.31, 95% HPD region: 1.24—1.39). Country-specific effects are apparent for the inequity in ownership based on gender (Figure 4A) and urban/rural residency (Figure 4B), and for the intercept (Figure 4—figure supplement 1); i.e., certain countries have greater (or lesser) inequities than the average effects for the 33 countries.

**TABLE 3:**
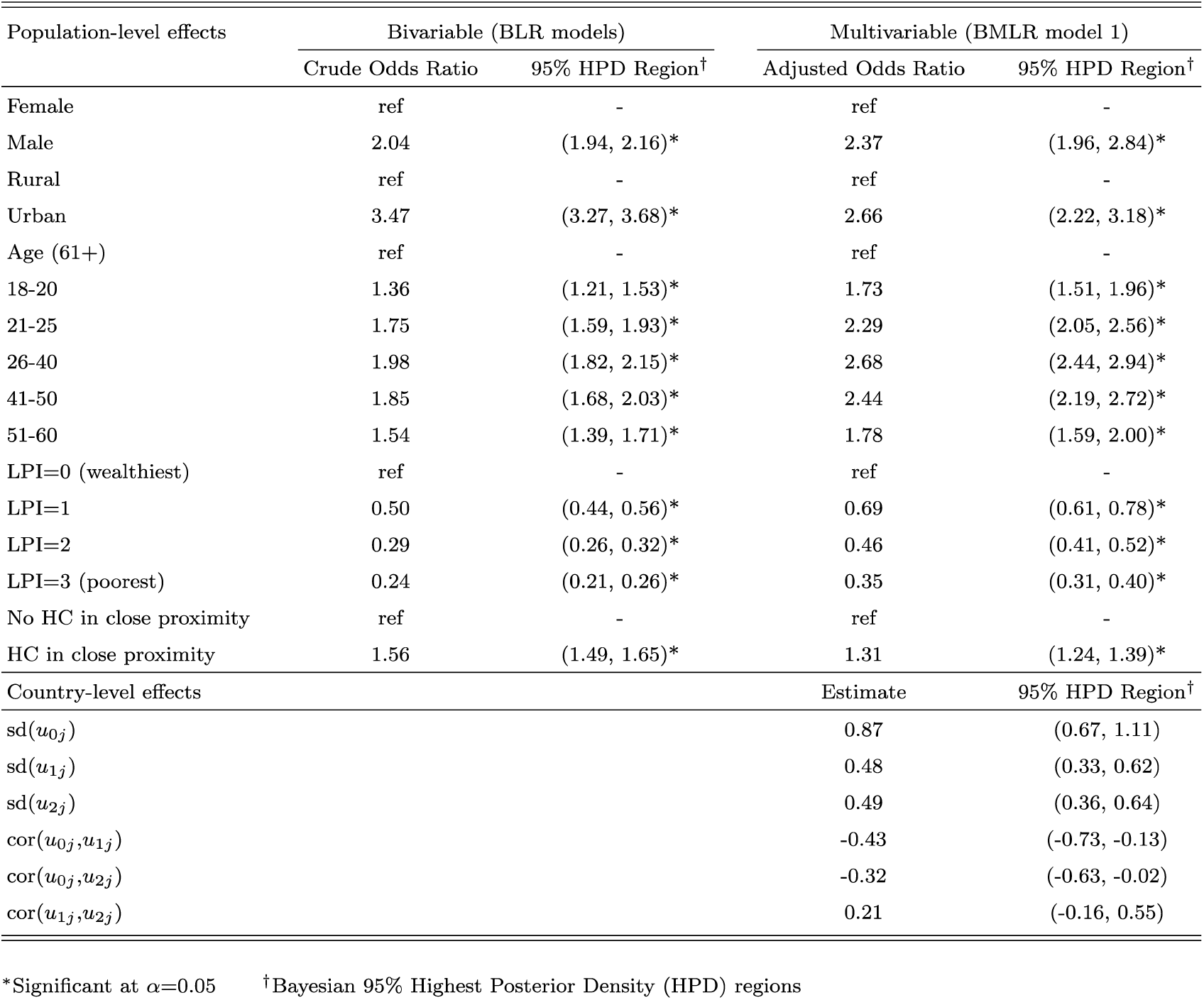
Fitted Bayesian models for determinants of mobile phone ownership in Africa

**Figure 4:**
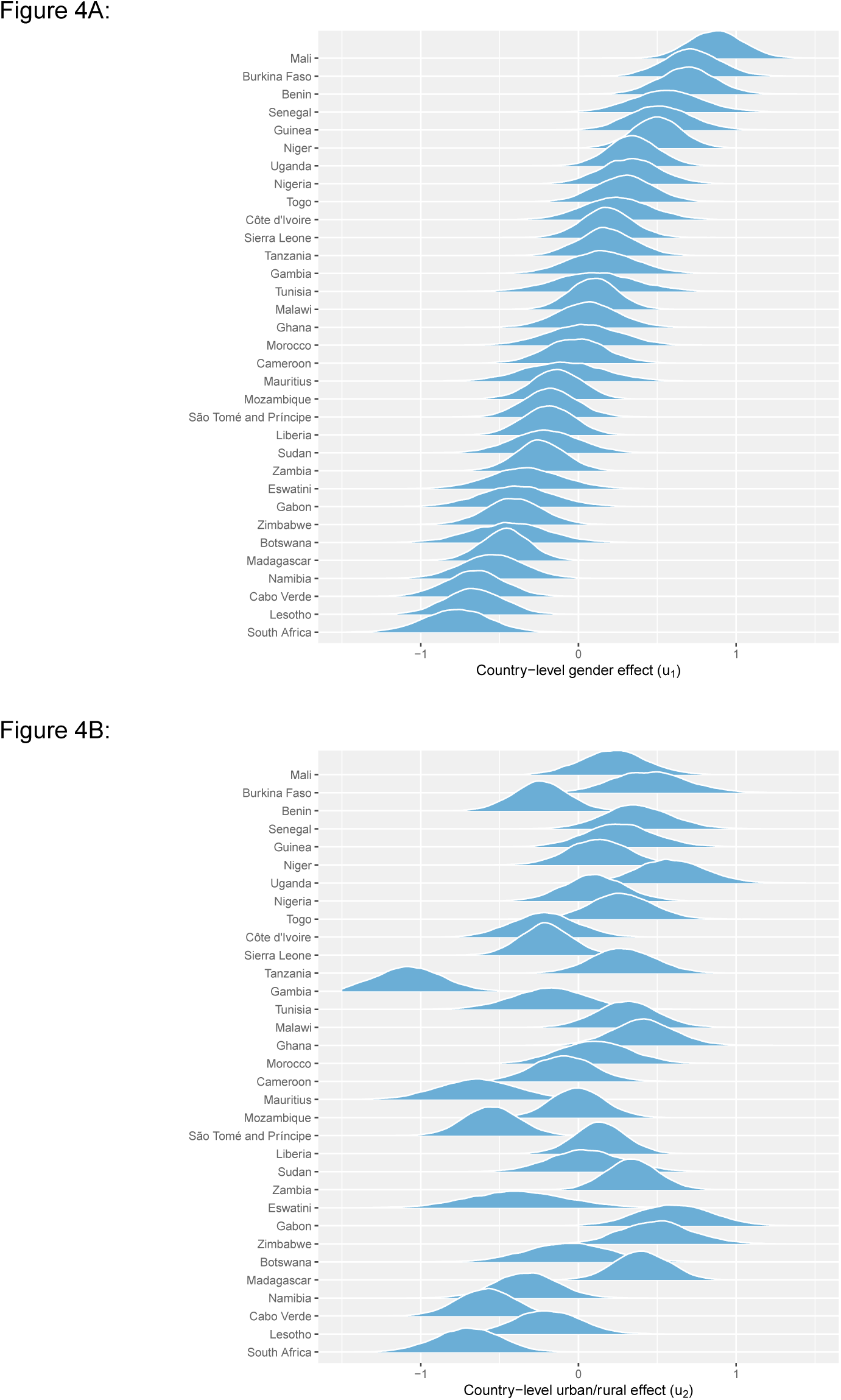
Country-level gender and urban-rural effects. **(A)** Posterior distributions of the country-level effect on mobile phone ownership of being male (compared to female), sorted by median. **(B)** Posterior distributions of the country-level effect of living in an urban area (compared to living in a rural area), in the same country order as (A). Both (A) and (B) are on the logit-scale – and should be viewed as country-specific adjustments to the population-level effects of (A) being male and (B) living in an urban area.

For mobile phone owners who own an SP, the results from the BLR and BMLR models are shown in Table 4. All ORs are significant different from 1 in both the bivariable and multivariable analysis. We found an interaction effect between gender and living in proximity to an HC: specifically, the effect of gender on owning an SP depends on proximity to an HC. Men who do not live in close proximity to an HC had higher odds of SP ownership (aOR: 1.50, 95% HPD region: 1.30—1.72) than women who do not live in close proximity (1; the baseline category). The gender difference is accentuated by proximity to an HC: men in close proximity to an HC had even larger odds of owning an SP (aOR: 1.92, 95% HPD region: 1.63—2.26) than women in close proximity (aOR: 1.15, 95% HPD region: 1.03—1.30). Urban residents (who own mobile phones) are nearly three times as likely as rural residents (who own mobile phones) to own SPs (aOR: 2.67, 95% HPD region: 2.33—3.10). Ownership of SPs (amongst owners of mobile phones) is most likely in 18 to 30 year olds, and decreases with age. The odds of ownership decrease with the LPI: the richest individuals (who own mobile phones) are approximately three times more likely than the poorest individuals (who own mobile phones) to own SPs (aOR: 2.86, 95% HPD region: 2.57—3.17). Country-specific effects are shown in Figure 4—figure supplements 2-F6). Notably, in many of the 33 countries, ownership of mobile phones is relatively high even amongst the poorest of the wealth classes; this holds true for both BPs and, more surprisingly, SPs (Figure 1—figure supplement 1). MCMC diagnostic plots and ROC curves for models 1 and 2 are provided in Figure 4—figure supplements 7-F8.

**TABLE 4:** Fitted Bayesian models for determinants of smartphone ownership among mobile phone owners in Africa

| Population-level effects | Bivariable (BLR models) |  | Multivariable (BMLR model 2) |  |
| --- | --- | --- | --- | --- |
|  | Crude Odds Ratio | 95% HPD Region<sup>†</sup> | Adjusted Odds Ratio | 95% HPD Region<sup>†</sup> |
| Female, no HC in close proximity | ref | - | ref | - |
| Female, HC in close proximity | 1.23 | (1.15, 1.32)* | 1.15 | (1.03, 1.30)* |
| Male, no HC in close proximity | 1.10 | (1.03, 1.18)* | 1.50 | (1.30, 1.72)* |
| Male, HC in close proximity | 1.54 | (1.44, 1.65)* | 1.92 | (1.63, 2.26)* |
| Rural | ref | - | ref | - |
| Urban | 2.95 | (2.81, 3.09)* | 2.67 | (2.33, 3.10)* |
| Age (61+) | ref | - | ref | - |
| 18-30 | 5.59 | (5.08, 6.16)* | 7.85 | (7.04, 8.77)* |
| 31-40 | 3.53 | (3.19, 3.90)* | 4.73 | (4.24, 5.30)* |
| 41-50 | 2.25 | (2.02, 2.50)* | 2.75 | (2.44, 3.09)* |
| 51-60 | 1.58 | (1.41, 1.77)* | 1.72 | (1.51, 1.96)* |
| LPI=0 (wealthiest) | ref | - | ref | - |
| LPI=1 | 0.61 | (0.57, 0.66)* | 0.67 | (0.62, 0.73)* |
| LPI=2 | 0.38 | (0.35, 0.40)* | 0.45 | (0.41, 0.49)* |
| LPI=3 (poorest) | 0.27 | (0.25, 0.29)* | 0.35 | (0.32, 0.39)* |
| Country-level effects |  |  | Estimate | 95% HPD Region<sup>†</sup> |
| $sd(u_{0j})$ | | | 0.85 | (0.67, 1.05) |
| $sd(u_{1j})$ | | | 0.39 | (0.28, 0.50) |
| $sd(u_{2j})$ | | | 0.21 | (0.08, 0.34) |
| $sd(u_{3j})$ | | | 0.31 | (0.18, 0.43) |
| $sd(u_{4j})$ | | | 0.41 | (0.28, 0.53) |
| $cor(u_{0j}, u_{1j})$ | | | -0.61 | (-0.81, -0.37) |
| $cor(u_{0j}, u_{2j})$ | | | -0.38 | (-0.77, 0.01) |
| $cor(u_{0j}, u_{3j})$ | | | -0.51 | (-0.80, -0.19) |
| $cor(u_{0j}, u_{4j})$ | | | -0.78 | (-0.93, -0.60) |
| $cor(u_{1j}, u_{2j})$ | | | 0.10 | (-0.38, 0.54) |
| $cor(u_{1j}, u_{3j})$ | | | 0.16 | (-0.24, 0.55) |
| $cor(u_{1j}, u_{4j})$ | | | 0.29 | (-0.06, 0.60) |
| $cor(u_{2j}, u_{3j})$ | | | -0.07 | (-0.59, 0.41) |
| $cor(u_{2j}, u_{4j})$ | | | 0.47 | (0.04, 0.82) |
| $cor(u_{3j}, u_{4j})$ | | | 0.61 | (0.3, 0.88) |

\*Significant at $\alpha=0.05$
<sup>†</sup>Bayesian 95% Highest Posterior Density (HPD) regions

## DISCUSSION

Considering all 33 African countries together, our results show that a fairly high proportion (82%) of individuals own a mobile phone, and ownership of either a BP or an SP is equally likely: however, we found considerable variability — amongst (and within) the 33 countries — in the proportion of individuals who own mobile phones. Furthermore, we found substantial inequities in ownership based on: gender (with men significantly more likely to own mobile phones than women), the urban-rural divide (with urban residents significantly more likely to own mobile phones than rural residents), and the LPI (wealthy individuals significantly more likely to own mobile phones than poor individuals); surprisingly, we found that some of the poorest individuals in all 33 countries own SPs. All of the inequities were exacerbated when ownership of an SP was considered in comparison to ownership of a BP. We also found that ownership increases with age, peaks in the age class 26 to 40 year olds, and then decreases. Due to the demographic structure of Africa, the majority of phone owners are under 30 years old. In this age category individuals are more likely to own an SP than a BP; older individuals are more likely to own a BP than an SP. Notably, we found that not all countries had gender inequities in ownership of mobile phones, but — essentially — all countries have substantial urban-rural inequities in ownership. In future work, we will identify explanatory factors underlying the inequities in phone ownership that we have found exist both between, and within, countries.

Our results have important implications for designing and rolling out mHealth interventions in Africa; mHealth interventions have the potential to increase the quality, reduce the cost and extend the reach of healthcare (Ag Ahmed et al., 2017; Manby et al., 2022; Onukwugha et al., 2022; Osei et al., 2020). Those who are most in need of mHealth interventions are those in rural areas who do not live in close proximity to an HC; these individuals will need to own mobile phones to access these mHealth interventions. However, our results show that these individuals are currently far less likely to own mobile phones than individuals who may have less need of mHealth interventions: i.e., individuals in urban areas who live in close proximity to an HC. Notably, our results demonstrate that mHealth interventions need to be designed to take age, geographic variation, and inequities (based on gender, urban/rural residency and wealth) in ownership of BPs and SPs into consideration. Previous studies have shown that it will also be critical to consider the impact of education and low levels of literacy on the ownership and usage of mobile phones, and hence on the design of mHealth interventions (Krönke, 2020). Our results suggest that, due to current levels of mobile phone ownership, it may only be possible to scale up mHealth interventions in a few countries (e.g., Botswana) (United Nations, 2021), but not in the vast majority of the 33 countries that we have analyzed. SPs are not yet widely used in many African countries; hence in the foreseeable future high-tech mHealth interventions based on SPs, rather than lower-tech interventions that only require BPs, will be hard to implement in the low-ownership countries that we have identified in our analyses. Taken together, our results demonstrate that before the promise of mHealth interventions can be reached in Africa, there is a need for a “digital transformation” to occur throughout the continent.

In 2019, the United Nations Broadband Commission for Sustainable Development proposed that there was a need for a “Digital Infrastructure Moonshot for Africa” (Broadband Commission, 2019). In 2020, the African Union (a continental body consisting of the member states that make up the countries of the African Continent) outlined specific goals that need to be met: “By 2030 all our people should be digitally empowered and able to access safely and securely to at least (6 megabyte per second) all the time where ever they live in the continent at an affordable price of no more than (1 cent [US dollars] per megabyte) through a smart device manufactured in the continent at the price of no more than (100 [US dollars]) to benefit from all basic e-services and content of which at least 30% is developed and hosted in Africa” (African Union, 2020). Since our analyses have revealed that only 40% of people in 33 of the 54 countries in Africa own SPs, these goals may currently be more aspirational than achievable. There are major infrastructural barriers — mainly in rural areas — that will need to be overcome: e.g., extending the electricity grid, increasing cell phone coverage, and expanding bandwidth (Broadband Commission, 2019; Krönke et al., 2022; Leo et al., 2015; World Health Organization, 2021b). There are also the inequities in ownership of SPs that we have identified in our analysis (gender, urban/rural residency, age, wealth) that will also need to be overcome in order to reach the goal of universal access to smart devices. Importantly, there will be a need to ensure sustainability: i.e., that owners of mobile phones can afford to pay for data and continue to utilize their devices. Notably, paying for broadband services in Africa is extremely expensive: in comparison to income, African countries have the highest prices worldwide (Broadband Commission, 2019).

Our study has several limitations. First, our analyses are only based on data from 33 out of 54 African countries. We recommend, when/if data become available, conducting the same analyses (as we have conducted here) for the 21 other countries. However, we believe that our qualitative results (i.e., ownership of mobile phones is fairly high, but ownership of SPs is relatively low, and substantial inequities in ownership exist) are likely to be generalizable to those 21 countries. Second, we have shown that individuals who live in closer proximity to HCs are more likely to own mobile phones. However, we have not demonstrated that owners of mobile phones have access to better healthcare than non-owners. Third, we have analyzed the most recent available Afrobarometer data (R7), collected in 2017-2018. We plan to analyze data from R8; these data are not yet available. We expect that this future analysis will show that levels of ownership will have increased over the past three to four years.

Mobile phone ownership is predicted to keep on growing in Africa, but to what degree this expansion will be in the ownership of BPs or in the ownership of SPs is unknown. Currently, the majority of Africans own a mobile phone, but less than half own an SP. Africa is becoming increasingly urbanized, and urban residents are more likely to be able to afford SPs than rural residents. However, increasing poverty levels throughout Africa — particularly in the African countries where the ownership of SPs is currently low — could limit the expansion in ownership of SPs; ownership of BPs could continue to increase. Some countries (e.g., Botswana) have developed specific plans for digital expansion (United Nations, 2021); their plans include strategies for overcoming the current barriers to ownership of smart devices and discussion of the economic, educational, agricultural, and health benefits that could result from such an expansion. Other country-specific plans will need to be developed. As the “digital transformation” of Africa continues, it will become critical to overcome the current urban-rural, gender, and wealth inequities in mobile phone ownership, particularly, the inequities in the ownership of SPs. If the digital devices needed for mHealth interventions are not equally available within the population (which we have found is the current situation), rolling out mHealth interventions in Africa is likely to propagate already existing inequities in access to healthcare.

## Data Availability

All data used in the paper is freely available at: https://afrobarometer.org/data/merged-data

## Data availability statement

All data used in the paper are freely available at: https://afrobarometer.org/data/merged-data. All code is freely available by emailing the corresponding author.

## Acknowledgements

We acknowledge Afrobarometer for making available all of the data that were used.

## List of figure supplements

**Figure 1—figure supplement 1.**
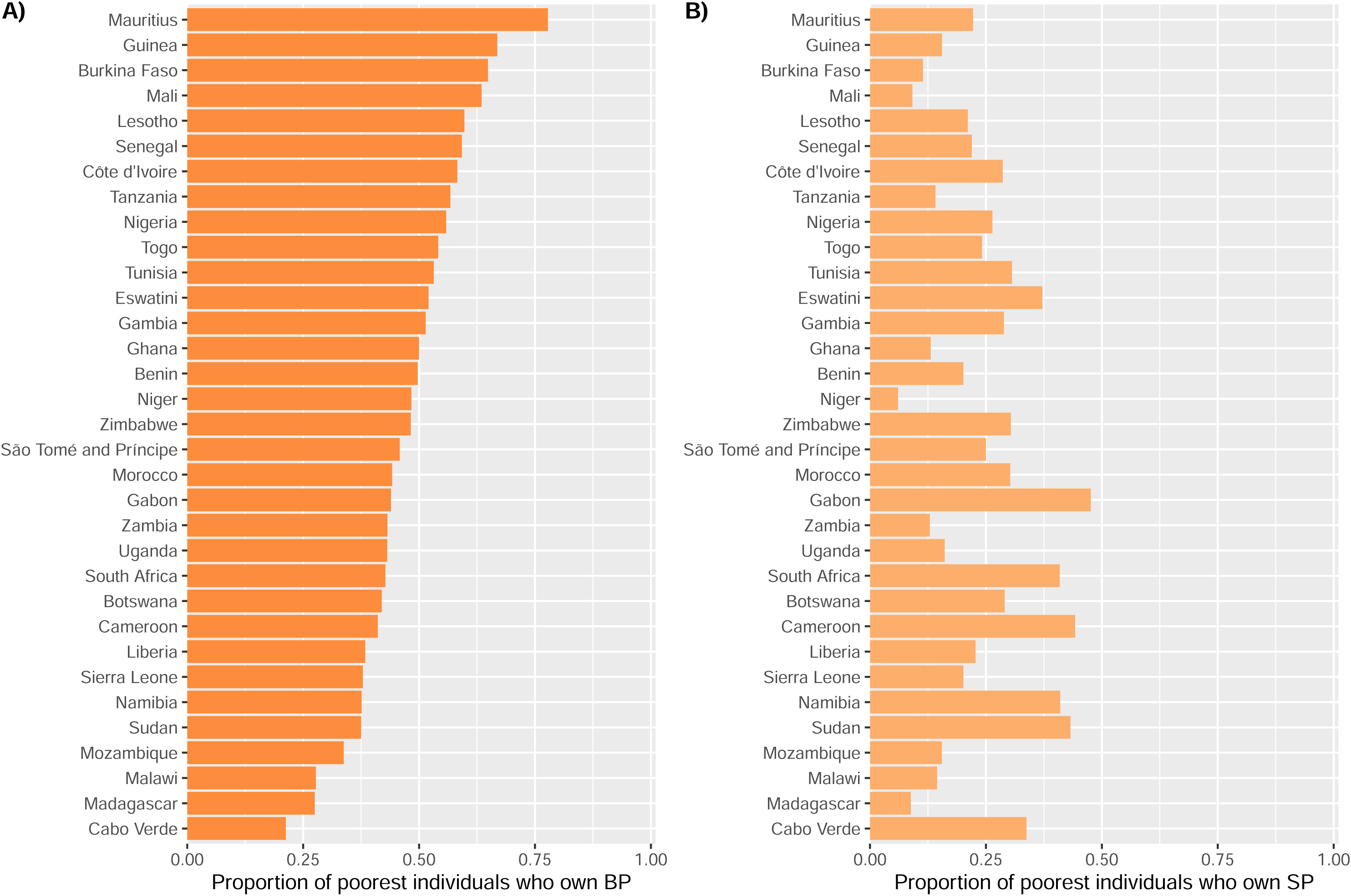
Phone ownership in the poorest individuals. **(A**) Country-level BP ownership in individuals with LPI=3. **(B)** Country-level SP ownership in individuals with LPI=3.

**Figure 4—figure supplement 1.**
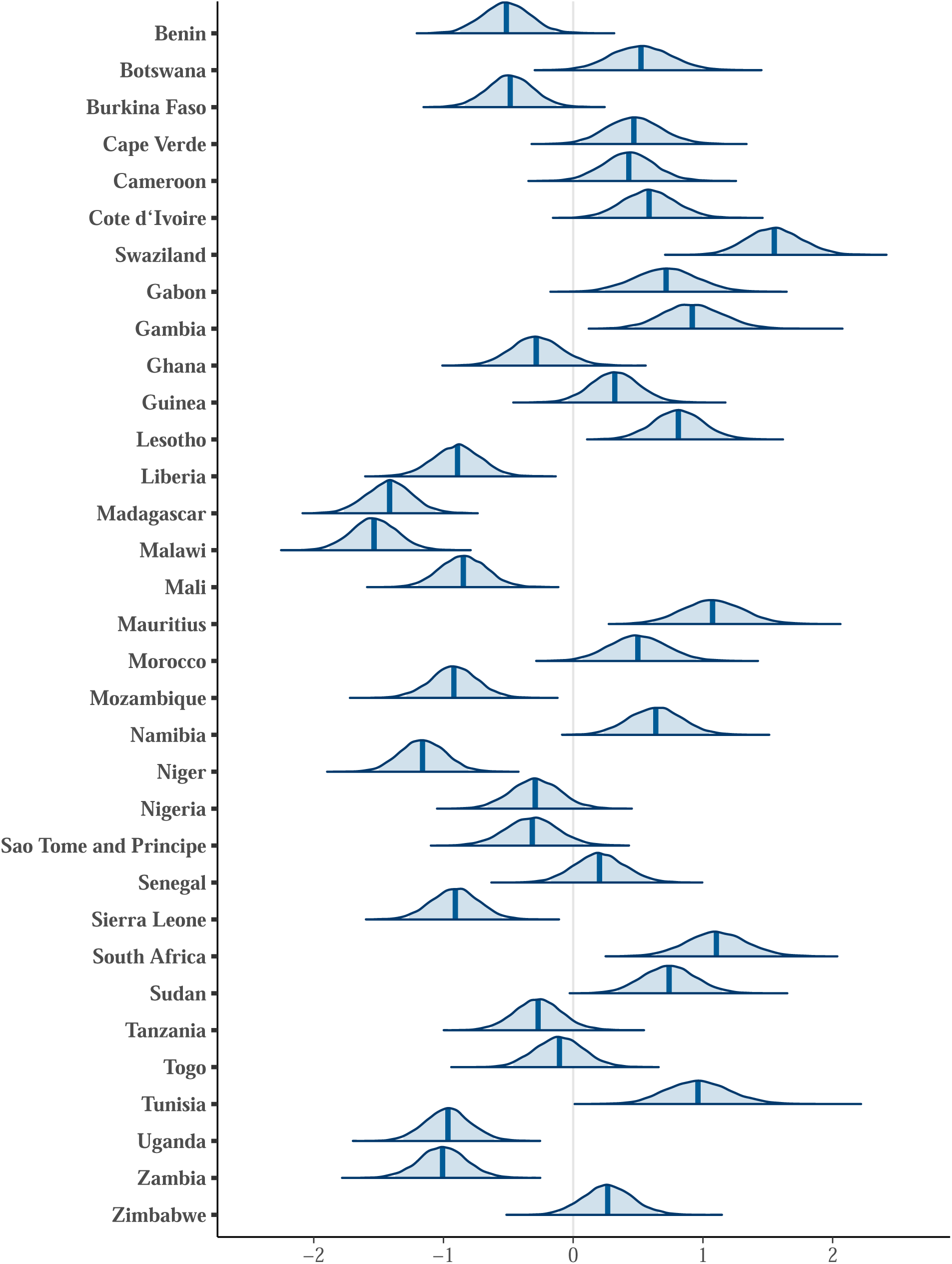
Model 1 posteriors – (country-level) intercept. Posterior distributions (medians and 95% HPD regions) of 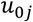, the country-level intercept, for country *j*.

**Figure 4—figure supplement 2.**
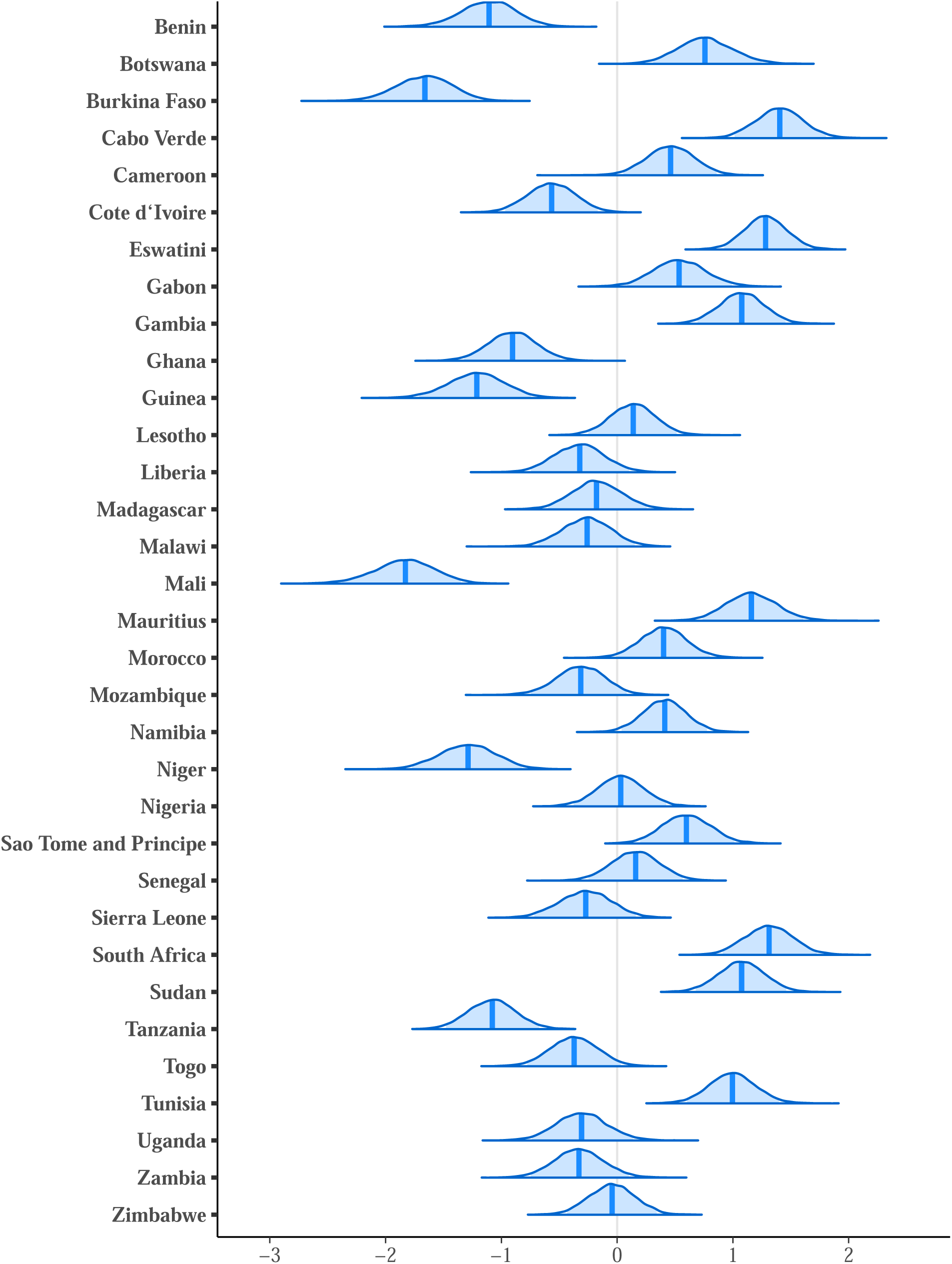
Model 2 posteriors – (country-level) intercept. Posterior distributions (medians and 95% HPD regions) of 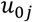 the country-level intercept, for country *j*.

**Figure 4—figure supplement 3.**
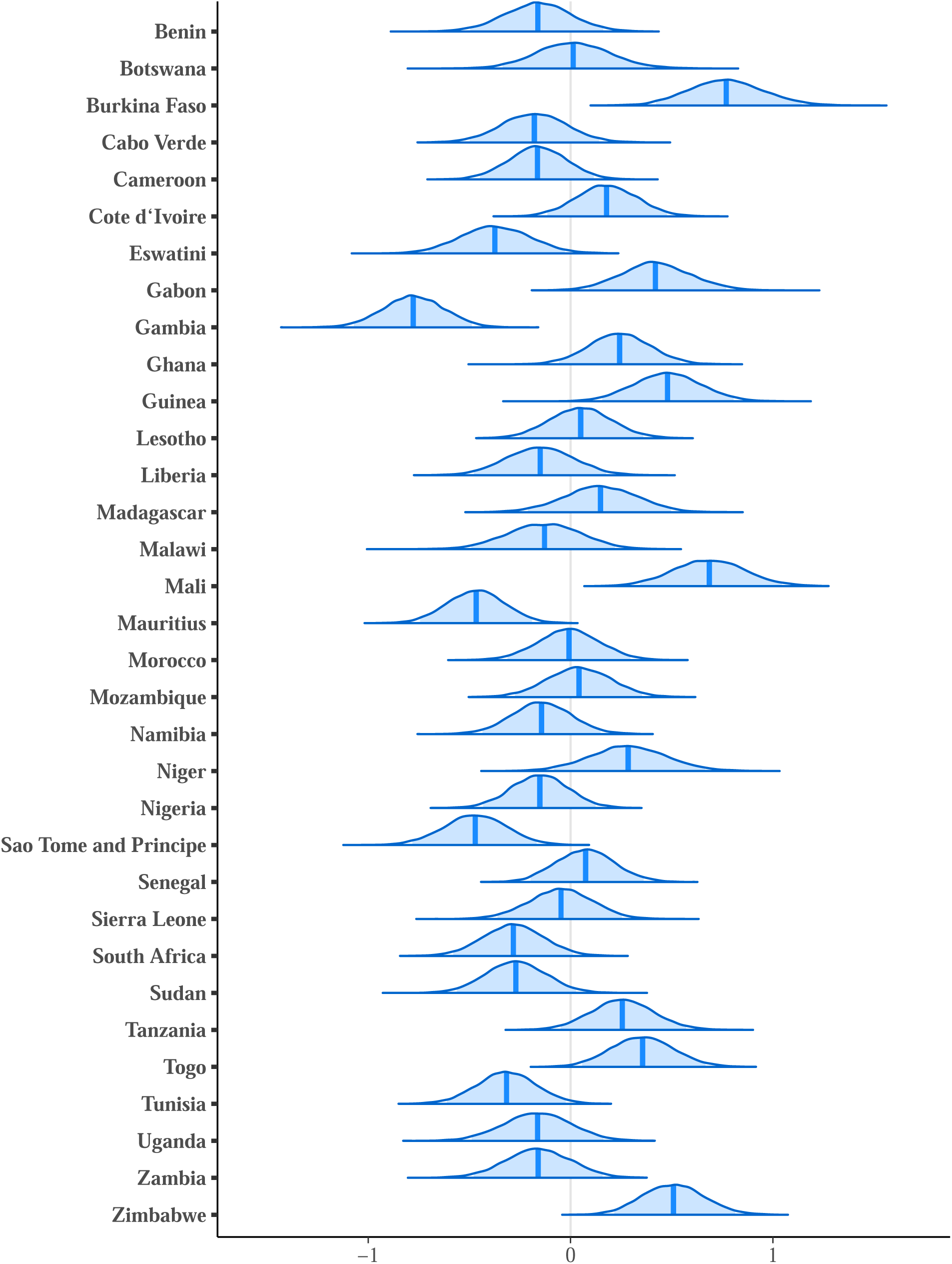
Model 2 posteriors – (country-level) urban/rural. Posterior distributions (medians and 95% HPD regions) of 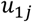, the country-level effect of living in an urban area, for country *j*.

**Figure 4—figure supplement 4.**
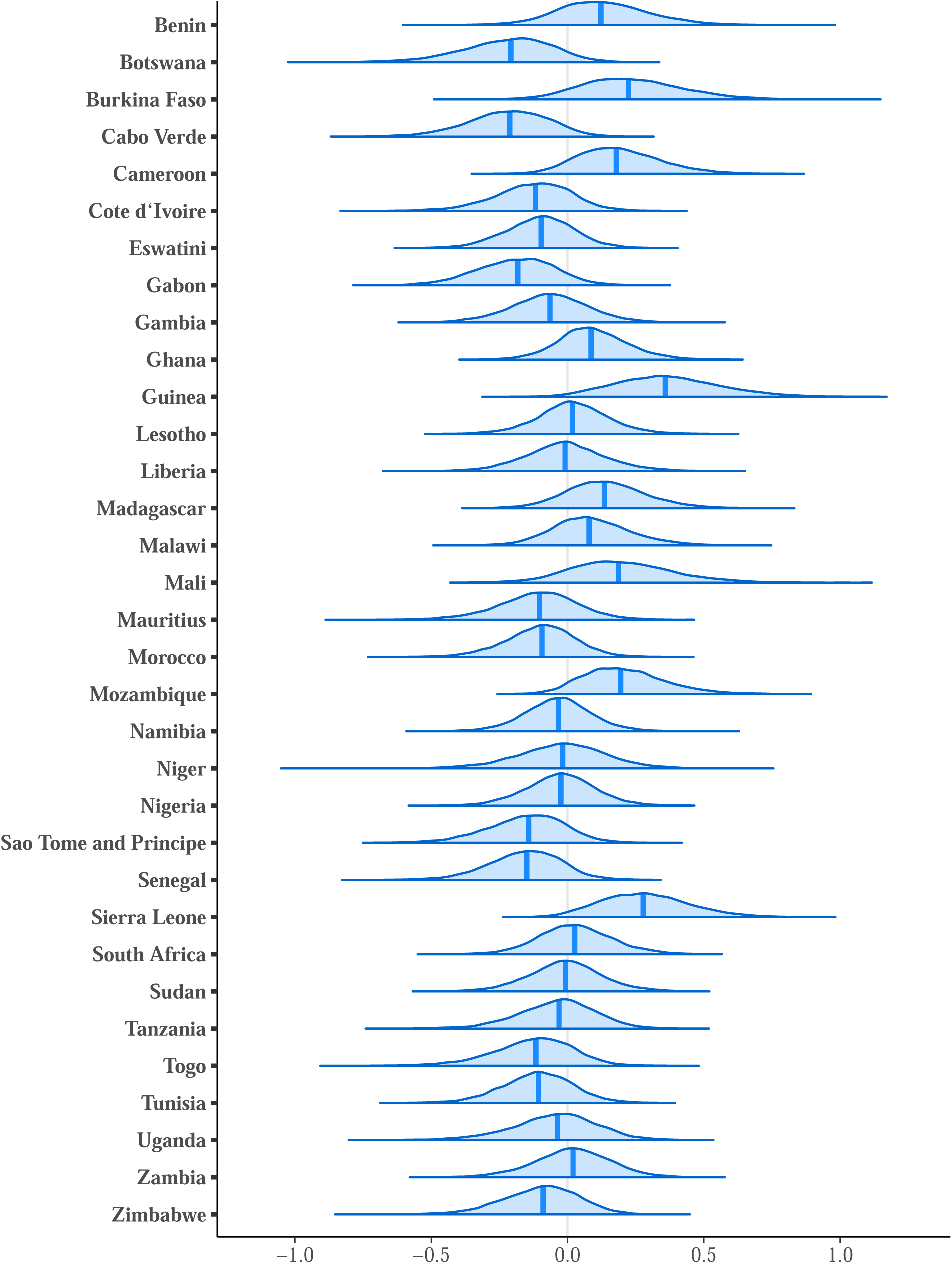
Model 2 posteriors – (country-level) gender and proximity. Posterior distributions (medians and 95% HPD regions) of 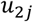, the country-level effect of being female in close proximity to an HC, for country *j*.

**Figure 4—figure supplement 5.**
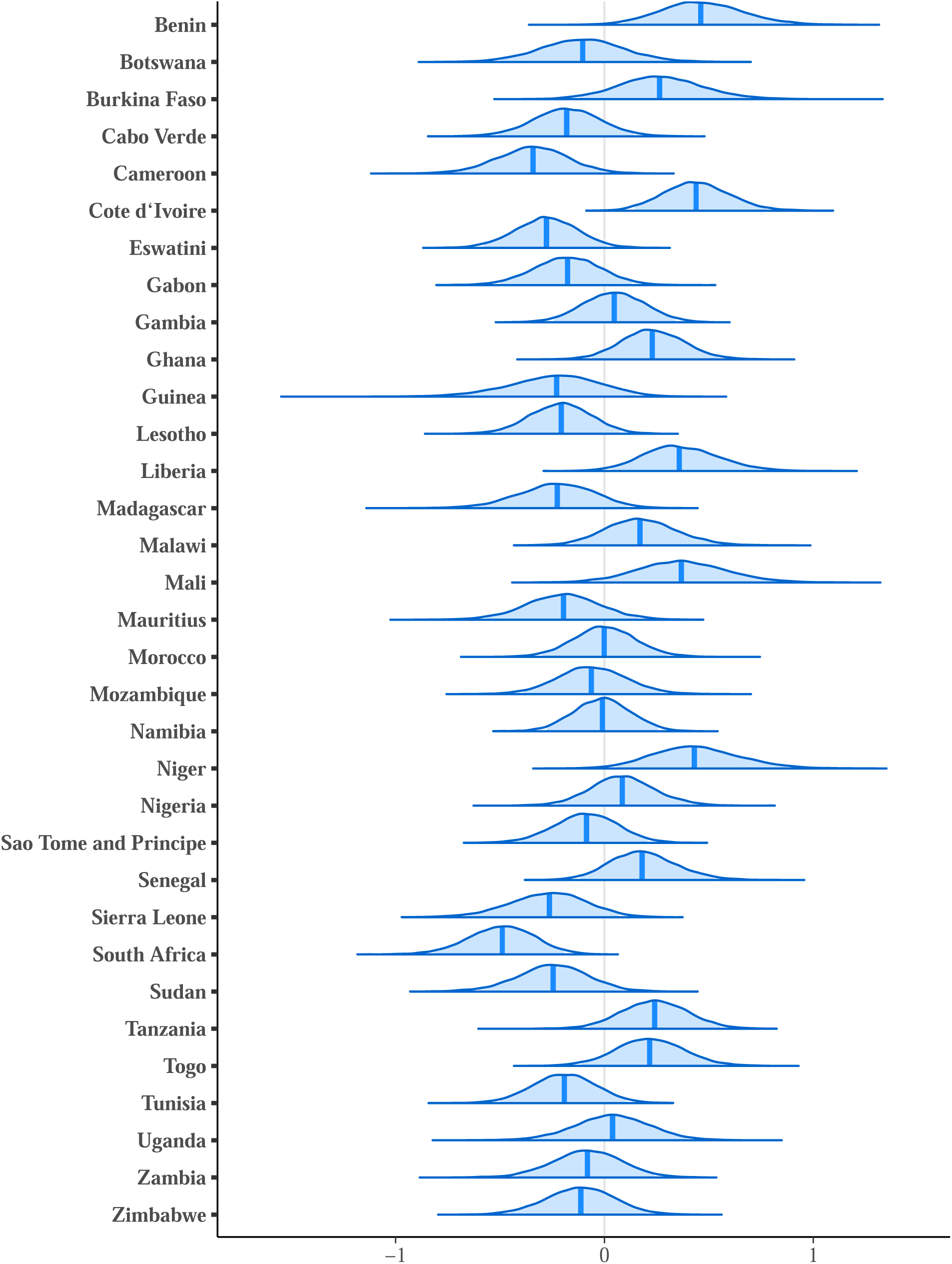
Model 2 posteriors – (country-level) gender and proximity. Posterior distributions (medians and 95% HPD regions) of 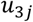, the country-level effect of being male not in close proximity to an HC, for country *j*.

**Figure 4—figure supplement 6.**
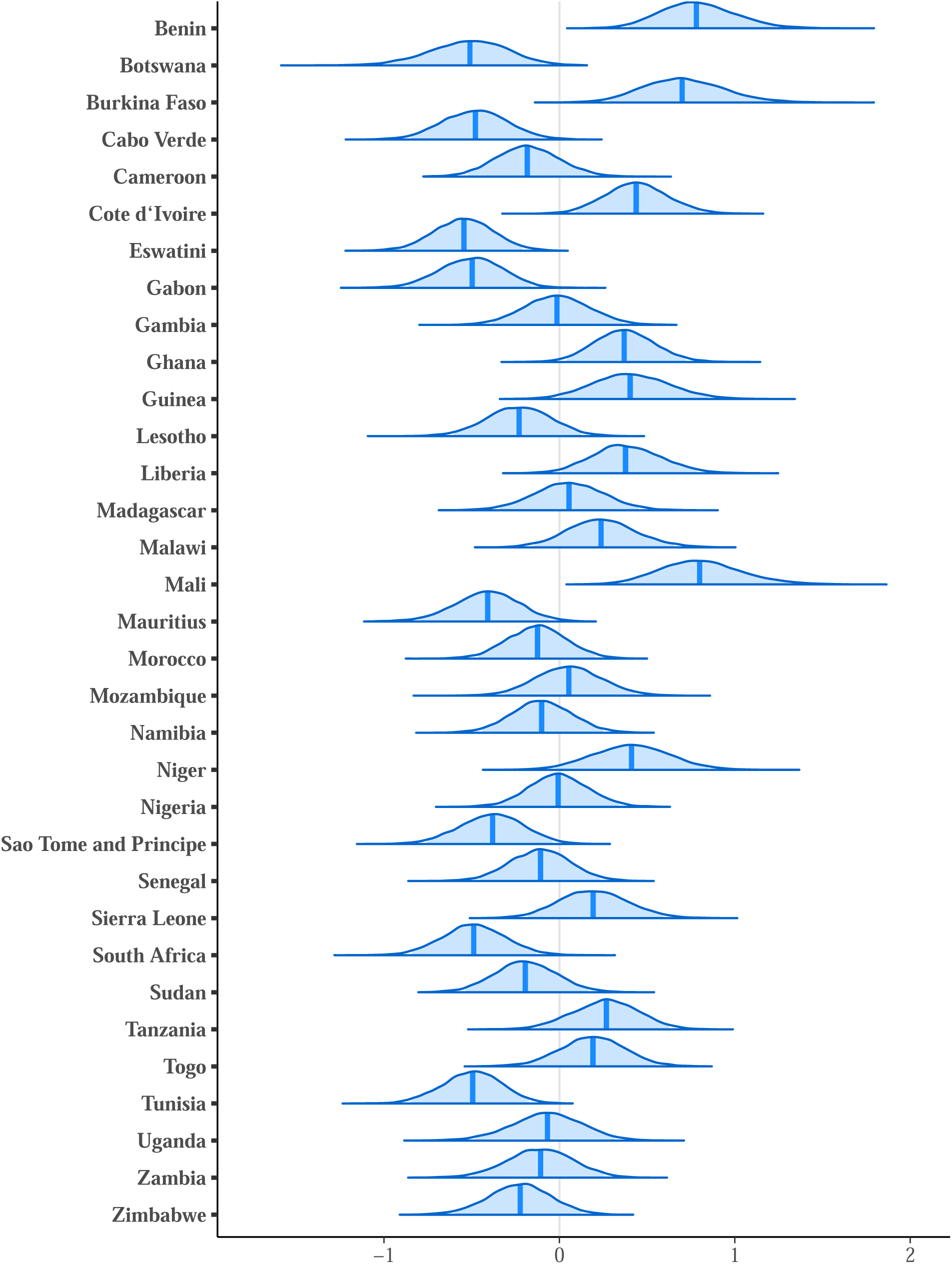
Model 2 posteriors – (country-level) gender and proximity. Posterior distributions (medians and 95% HPD regions) of 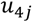, the country-level effect of being male in close proximity to an HC, for country *j*.

**Figure 4—figure supplement 7.**
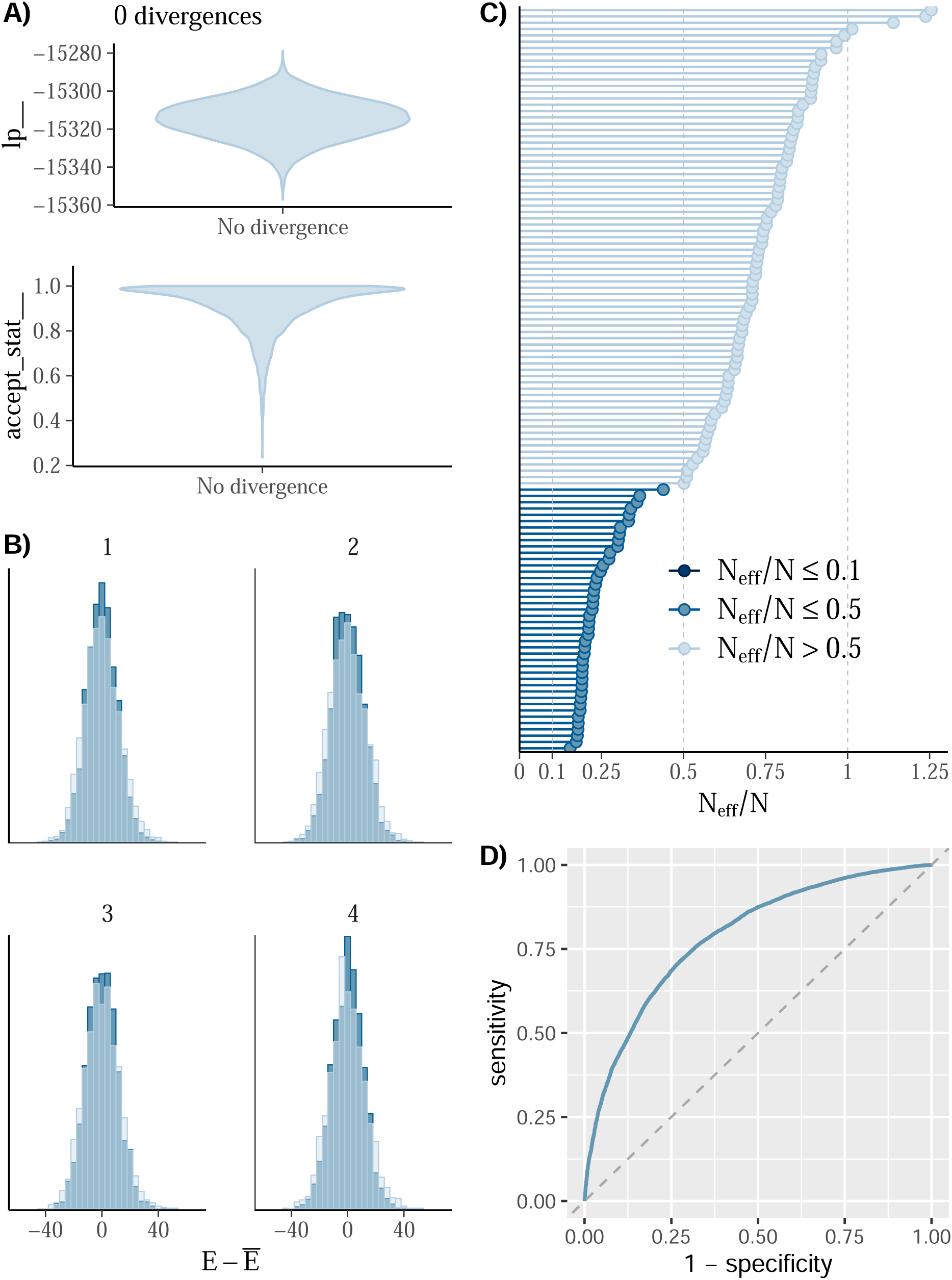
Model 1 MCMC diagnostics and ROC curve. **(A)** Violin plots of the log posterior (top) and No-U-Turn Sampling (NUTS) acceptance statistic (bottom). There are no divergent transitions. To further assess chain convergence, we used 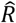, the potential scale reduction factor (Gelman & Rubin, 1992). All values of 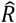 are nearly equal to 1 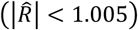, indicating all chains have converged. **(B)** NUTS energy plot (Betancourt, 2017) for all 4 chains. The transition distribution (darker histogram in each plot) and target distribution (lighter histogram in each plot) are moderately well aligned, indicating efficient sampling. **(C)** The effective sample size *N*_*eff*_ from the posterior distribution of each parameter was used to gauge the level of autocorrelation in each sample. *N*_*eff*_/*N* > 0.1 for all model parameters indicating independent draws within each sample. **(D)** ROC curve used to assess the diagnostic capability of fitted BMLR model. The Area Under the Curve (AUC) was 0.79 indicating good predictive accuracy.

**Figure 4—figure supplement 8.**
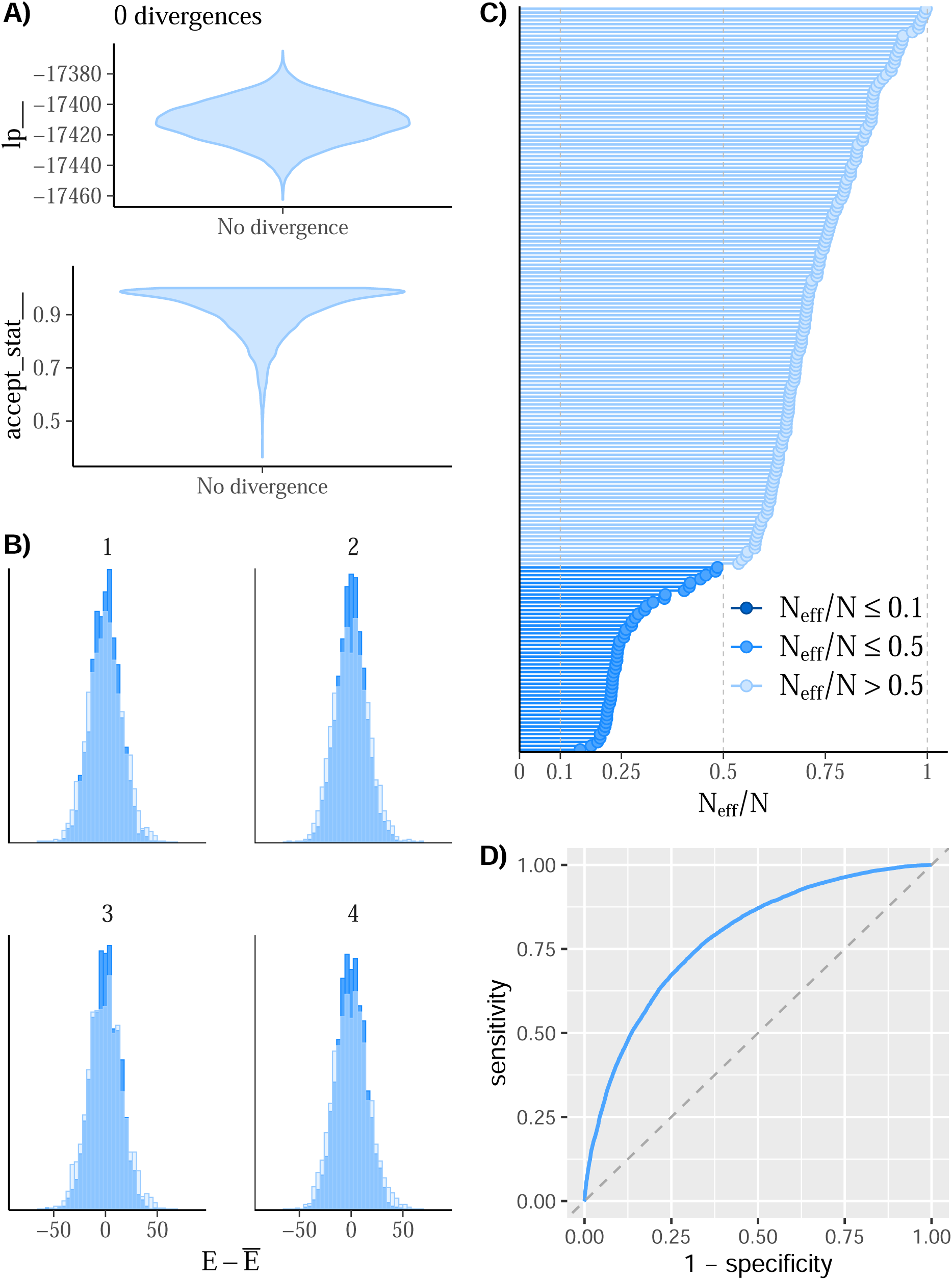
Model 2 MCMC diagnostics and ROC curve. **(A)** Violin plots of the log posterior (top) and NUTS acceptance statistic (bottom). There are no divergent transitions. Additionally, all values of 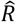 are nearly equal to 1 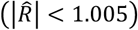, indicating all chains have converged. **(B)** NUTS energy plot for all 4 chains. The transition distribution (darker histogram in each plot) and target distribution (lighter histogram in each plot) are moderately well aligned, indicating efficient sampling. **(C)** *N*_*eff*_/*N* > 0.1 for all model parameters indicating independent draws within each sample. **(D)** ROC curve used to assess the diagnostic capability of fitted BMLR model. The AUC was 0.78 indicating good predictive accuracy.

